# Plasma biomarkers of neurodegeneration in patients with Parkinson’s disease and dementia with Lewy bodies and at-risk individuals of Lewy body disease in the NaT-PROBE cohort

**DOI:** 10.1101/2023.12.03.23299326

**Authors:** Keita Hiraga, Makoto Hattori, Yuki Satake, Daigo Tamakoshi, Taiki Fukushima, Takashi Uematsu, Takashi Tsuboi, Maki Sato, Katsunori Yokoi, Keisuke Suzuki, Yutaka Arahata, Yukihiko Washimi, Akihiro Hori, Masayuki Yamamoto, Hideaki Shimizu, Masakazu Wakai, Harutsugu Tatebe, Takahiko Tokuda, Akinori Nakamura, Shumpei Niida, Masahisa Katsuno

**Author notes:** Corresponding author: Masahisa Katsuno, MD, PhD, Department of Neurology, Nagoya University Graduate School of Medicine 65 Tsurumai-cho, Showa-ku, Nagoya, Japan, 466-8550. **E-mails:** (K.H.); (M.H.); (Y.S.); (D.T.); (T.F.); (T.U.); (T.Ts.); (M.S.); (K.Y.); (K.S.); (Y.A.); (Y.W.); (A.H.); (M.Y.); (H.S.); (M.W.); (H.T.); (T.To.); (A.N.); (S.N.); (M.K.).

## Abstract

**Background:** Comorbid Alzheimer’s disease (AD) neuropathology is common in Lewy body disease (LBD); however, AD comorbidity in the prodromal phase of LBD remains unclarified. This study investigated AD comorbidity in the prodromal and symptomatic phases of LBD by analysing plasma biomarkers in patients with Parkinson’s disease (PD) and dementia with Lewy bodies (DLB) and at-risk individuals of LBD (NaT-PROBE cohort).

**Methods:** Patients with PD (PD group, n=84) and DLB (DLB group, n=16) and individuals with LBD with ≥2 (high-risk group, n=82) and without (low-risk group, n=37) prodromal symptoms were enrolled. Plasma amyloid-beta (Aβ) composite was measured using immunoprecipitation-mass spectrometry assays. Plasma phosphorylated tau 181 (p-tau181), neurofilament light chain (NfL), and alpha-synuclein (aSyn) were measured using a single-molecule array.

**Results:** p-tau181 levels were higher in the PD and DLB groups than in the low-risk group. Aβ composite level was higher in the DLB group than in the high-risk group. AD-related biomarker levels were not elevated in the high-risk group. NfL levels were higher in the high-risk, PD, and DLB groups than in the low-risk group. In the PD group, Aβ composite was associated with cognitive function, p-tau181 with motor function and non-motor symptoms, and NfL with cognitive and motor functions and non-motor symptoms. In the high-risk group, NfL was associated with metaiodobenzylguanidine scintigraphy abnormalities.

**Conclusions:** The PD and DLB groups exhibited comorbid AD neuropathology, though not in the prodromal phase. Elevated plasma NfL levels, even without elevated AD-related plasma biomarker levels, may indicate aSyn-induced neurodegeneration in the prodromal phase of LBD.

*What is already known on this topic:* Comorbid Alzheimer’s disease (AD) neuropathology is common in Parkinson’s disease (PD) dementia and dementia with Lewy bodies (DLB); however, AD comorbidity in the prodromal phase of Lewy body disease (LBD) is yet to be clarified.

*What this study adds:* Four plasma biomarkers (amyloid-beta (Aβ) composite, phosphorylated tau 181 (p-tau181), neurofilament light chain (NfL), and alpha-synuclein (aSyn)) were measured in patients with PD and DLB and high– and low-risk individuals with ≥2 and without LBD prodromal symptoms. Increased plasma levels of p-tau181 indicated AD comorbidity in patients with PD and DLB, while no elevation of AD-related biomarkers was found in the high-risk individuals. Plasma NfL levels were higher in the high-risk, PD, and DLB groups than in the low-risk group and were associated with a higher rate of abnormalities in metaiodobenzylguanidine (MIBG) scintigraphy in the high-risk group.

*How this study might affect research, practice or policy:* This study demonstrated that comorbid AD neuropathology exists in the symptomatic phase of LBD, though not in its prodromal phase. Plasma p-tau181 may be more sensitive for detecting comorbid AD neuropathology than plasma Aβ composite. Elevated plasma NfL levels may indicate aSyn-induced neurodegeneration in the prodromal phase of LBD.

## INTRODUCTION

Lewy body disease (LBD) includes Parkinson’s disease (PD) and dementia with Lewy bodies (DLB), which are neurodegenerative disorders associated with intra-neuronal alpha-synuclein (aSyn) accumulation. Prodromal symptoms of LBD, including dysautonomia, hyposmia, and rapid eye movement sleep behaviour disorder (RBD), precede the onset of motor or cognitive dysfunction by 10–20 years and are considered essential for the pre-onset risk assessment of LBD development.[1]

In our previous high-risk cohort study for LBD, we found that 5.7% of healthy participants aged ≥50 years had ≥2 prodromal symptoms; we defined these participants as high-risk individuals.[2] These participants had mild cognitive decline and hyposmia compared with low-risk participants with no prodromal symptoms. Approximately one-third of the high-risk individuals had defects in dopamine transporter (DaT) single-photon-emission computed tomography (SPECT) or cardiac metaiodobenzylguanidine (MIBG) scintigraphy, and the prevalence of abnormalities on DaT-SPECT was 4 times higher in the high-risk individuals than in the low-risk individuals.[3]

In PD and DLB, limbic and neocortical aSyn pathology is associated with dementia; in addition, previous postmortem brain studies demonstrated that comorbid Alzheimer’s disease (AD) neuropathology is associated with the progression of cognitive impairment. More than 70% of patients with DLB and approximately 50% of patients with PD dementia (PDD) have comorbid AD neuropathology.[4, 5] Recently, AD-related plasma biomarkers, such as amyloid-beta (Aβ) composite (combination biomarker of amyloid-beta precursor protein (APP)_669-711_/Aβ_1-42_ and Aβ_1-40_/Aβ_1-42_ ratios) and phosphorylated tau 181 (p-tau181), have garnered attention.[6, 7] In addition, plasma neurofilament light chain (NfL) is regarded as a reliable biomarker for various neurodegenerative diseases.[8] Although recent studies have examined AD-related plasma biomarkers in patients with PD and DLB [9, 10] and plasma NfL in patients with idiopathic RBD,[11, 12] a detailed study on AD comorbidity in the prodromal phase of LBD is lacking.

Therefore, this study measured and analysed four plasma biomarkers, Aβ composite, p-tau181, NfL, and aSyn, in high-risk and low-risk individuals, participating in the NaT-PROBE study, as well as in patients with PD and DLB.

## METHODS

### Study design and participants

The Nagoya-Takayama preclinical/prodromal Lewy body disease (NaT-PROBE) study is a prospective, longitudinal, multi-centre, community-based cohort study coordinated by the Nagoya University School of Medicine. Between March 2017 and January 2023, healthy individuals aged ≥50 years who visited the Kumiai Kosei Hospital, Daido Clinic, or Chutoen General Medical Center, in Japan, for their annual health checkup have been surveyed using the following questionnaires: the Japanese version of the Scale for Outcomes in Parkinson’s disease for Autonomic Symptoms (SCOPA-AUT); the Self-administered Odor Question (SAOQ); the RBD screening scale (RBDSQ); the Beck Depression Inventory-Second Edition (BDI-II); the Epworth Sleepiness Scale (ESS); and the Physical Activity Scale for the Elderly (PASE).[3] Based on the results of our previous study,[2] 82 and 37 consecutive participants who had ≥2 abnormal scores (high-risk group) and no abnormalities (low-risk group), respectively, in the SCOPA-AUT, SAOQ, and RBDSQ scales were enrolled in the present study. The cut-off value for identifying the high-risk group was 10, 90.0%, and 5 for the SCOPA-AUT, SAOQ, and RBDSQ scales, respectively.[2]

In addition, patients with PD and DLB who visited Nagoya University Hospital, Kumiai Kosei Hospital, and the National Center for Geriatrics and Gerontology between March 2017 and January 2023 were evaluated. Among these, 84 patients with PD, who met the United Kingdom Parkinson’s Disease Society Brain Bank Diagnostic Criteria,[13] and 14 patients with DLB, who met the diagnostic criteria of the fourth report of the DLB consortium,[14] were enrolled in the present study.

Comprehensive evaluations, including cognitive and motor function assessments, questionnaire surveys, and blood sampling were conducted for all participants. Additionally, DaT-SPECT and cardiac MIBG scintigraphy were performed for all high– and low-risk participants.

### Cognitive and motor function examination

The Japanese version of the Montreal Cognitive Assessment (MoCA-J), the Stroop test, and the line orientation test were used to assess the general cognitive, frontal lobe, and visuospatial cognitive functions, respectively. Patients with PD with MoCA-J ≥26 and <26 were classified as cognitively normal (PD-CN) and cognitively impaired (PD-CI), respectively, according to a previously proposed criteria.[15] The Movement Disorder Society-Unified Parkinson’s Disease Rating Scale (MDS-UPDRS) was scored by neurologists who were certified MDS-UPDRS evaluators for assessing PD-related motor and non-motor symptoms. Rigidity (3.3), tremor (3.15–3.18), bradykinesia (3.2, 3.4–3.8, and 3.14), and axial signs (3.1 and 3.9–3.13) scores were extracted from the MDS-UPDRS III for further analysis. Levodopa equivalent daily dose (LEDD) was calculated as previously described.[16]

### Questionnaires on motor and non-motor symptoms

The SCOPA-AUT (Japanese version), SAOQ, RBDSQ, BDI-II, ESS, Parkinson’s Disease Questionnaire-39 (PDQ-39), and Questionnaire for Impulsive-Compulsive Disorders in Parkinson’s Disease (QUIP) were used to evaluate autonomic dysfunction, olfactory dysfunction, RBD, depressive symptoms excessive daytime sleepiness, PD-specific health-related quality of life, and impulse control disorder, respectively.

All the aforementioned questionnaires were validated for self-administration in a Japanese population.[17–23]

### Imaging tests

DaT-SPECT imaging with (^123^I)FP-CIT and cardiac (^123^I)MIBG scintigraphy (^123^I-MIBG) were performed to detect presynaptic dopamine neuronal dysfunction and to assess postganglionic cardiac autonomic denervation, respectively. DaT-SPECT and MIBG scintigraphy were measured as previously described.[3] DaT-SPECT was considered abnormal when decreased DaT-SPECT Specific Binding Ratio (SBR) or abnormal visual findings were found. The reference values of Japanese volunteers were used to evaluate the decrease in DaT SPECT SBR.[24] MIBG was considered abnormal when early or delayed H/M ratios were <2.2 or <2.2, respectively.[25]

### Sample collection and plasma biomarker measurements

Plasma samples, collected in EDTA-2Na-containing tubes, were centrifuged for 10 min at 1200 or 3000 × g, aliquoted, and immediately stored at −80 ℃. Plasma Aβ composite was measured via immunoprecipitation-mass spectrometry (IP-MS) assays as previously described.[6] Plasma p-tau181, NfL, and aSyn were measured by a single-molecule array (Simoa) using pTau-181 Advantage V2 Kit, NF-light Advantage V2 Kit, and Alpha-Synuclein Discovery Kit (Quanterix, Billerica, MA, USA).

The cut-off value for plasma Aβ composite was set at 0.376, based on previous studies.[6, 26] The plasma p-tau181 and NfL levels were log-transformed with base 10 to approximate a normal distribution, and the 95% Confidence Interval (CI) upper limit of the low-risk participants without abnormal plasma Aβ composite and DaT-SPECT and MIBG imaging was used for cut-off values (log_10_ (p-tau181), 0.374; log_10_ (NfL), 1.65). These cut-off values were used to determine the AT(N) profile[27] (A−, Aβ composite <0.376; A+, Aβ composite ≥0.376; T−, log_10_ (p-tau181) <0.374; T+, log_10_ (p-tau181) ≥0.374; N−, log_10_ (NfL) <1.65; N+, log_10_ (NfL) ≥1.65).

### Statistical analyses

All data represented the mean (standard deviation), unless otherwise stated. Since aSyn is abundant in red blood cells,[28] the aSyn/Hb ratio, corrected using haemoglobin levels, was used in the analysis. The demographic scores of the low-risk, high-risk, PD, and DLB groups were compared using a parametric one-way analysis of variance (ANOVA), followed by Tukey’s test. Between-group categorical variables were compared using Fisher’s exact test. The Benjamini–Hochberg method was used for multiple comparisons. The clinical scores and plasma biomarkers of the low-risk, high-risk, PD, and DLB groups were compared using the analysis of covariance (ANCOVA) adjusted for age and sex, followed by Tukey’s test using the Benjamini–Hochberg method. When comparing A+ and A−, T+ and T−, and N+ and N− among the patients with PD or high-risk participants, Student’s *t*-test was used for the demographic scores, and ANCOVA adjusted for age and sex was used for the clinical scores and plasma biomarkers. Pearson’s correlation coefficient was used to determine the relationship between plasma biomarkers and age. Age-adjusted Pearson’s partial correlation coefficient was used to determine the relationships between the plasma biomarkers and between the plasma biomarkers and each clinical score.

p values <0.05 were considered as statistically significant. Correlation coefficients (*r*) were interpreted as follows: >0.8, ‘very strong’; 0.5–0.8, ‘moderately strong’; and 0.3–0.5, ‘weak’. All statistical analyses were performed using R version 4.2.0, R Foundation for Statistical Computing, Vienna, Austria (https://www.R-project.org/). Figures were generated using the R package *ggplot2*.

## RESULTS

### Participant characteristics

There were more male participants in the low– and high-risk groups than in the PD and DLB groups. The PD and DLB group participants were significantly older than the low– and high-risk group participants. Among the high-risk participants, 36.6% had abnormalities in either DaT or MIBG, consistent with the findings in our previous study.[3] All patients with PD and DLB who underwent DaT or MIBG before the study inclusion exhibited these abnormalities. The PD and DLB group participants, though not the high-risk group participants, had worse scores of MoCA-J compared with the low-risk group participants. Two patients with PD and three with DLB could not complete the Stroop test, and one patient with DLB could not complete the line orientation test. The average Hoehn and Yahr Scale score was similar between the PD and DLB groups. The PD and DLB group participants, though not those in the high-risk group, had worse MDS-UPDRS III scores. The high-risk participants who were selected based on the SCOPA-AUT, SAOQ, and RBDSQ scores had worse BDI-II, ESS, PDQ-39, and QUIP scores than the low-risk participants. The PD and DLB group participants had worse scores on these questionnaires as well, except for QUIP (Table 1).

**Table 1.**
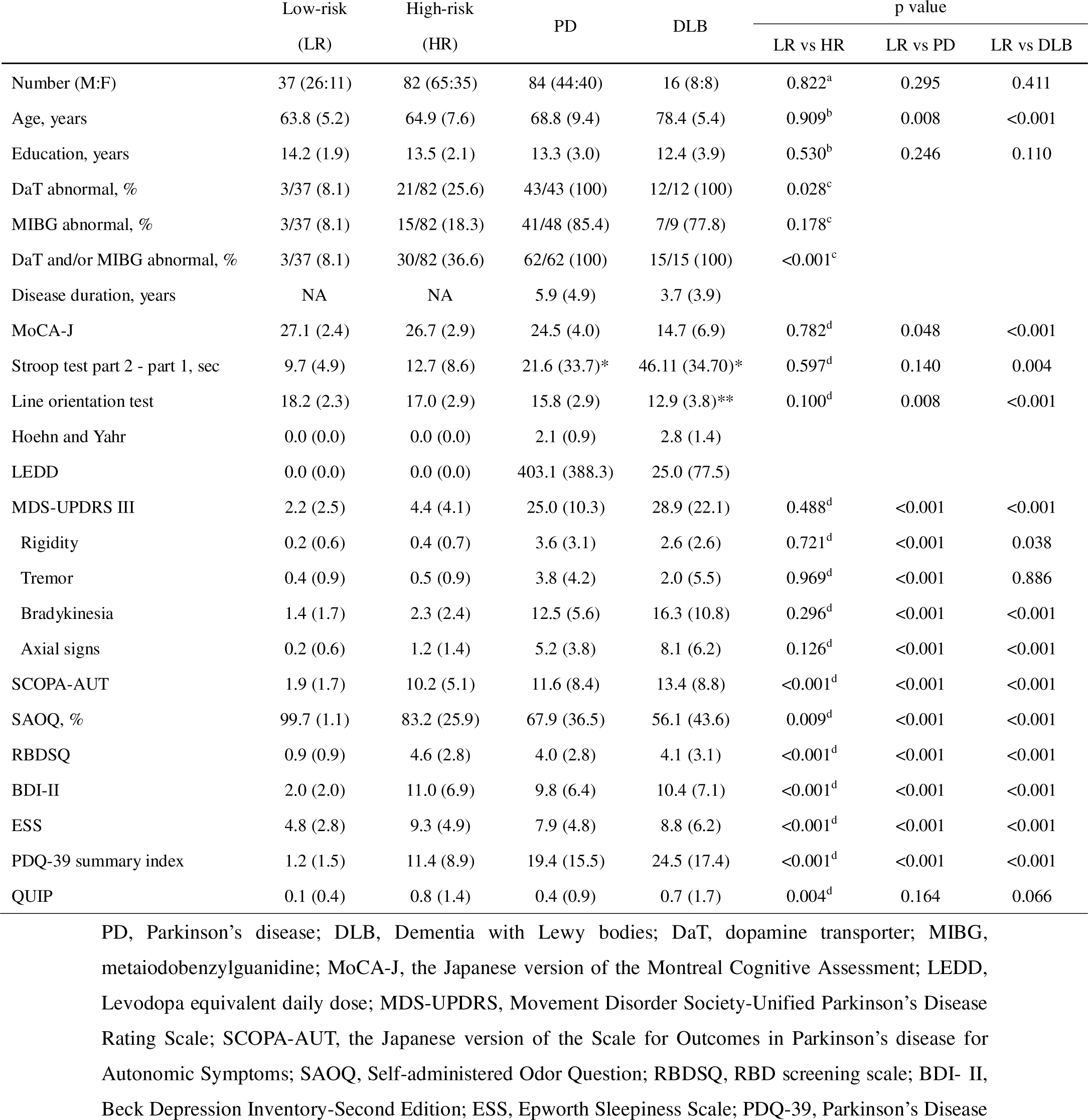
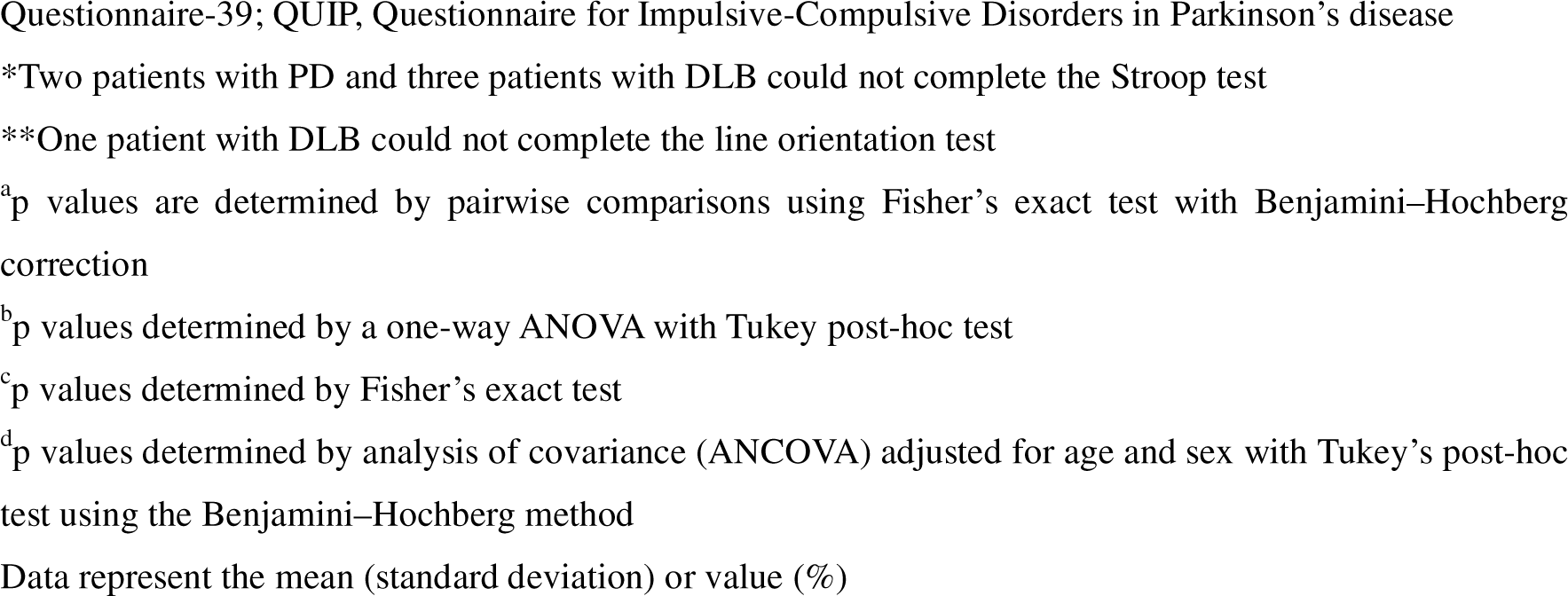
Background characteristics of the participants.

|  | Low-risk<br>(LR) | High-risk<br>(HR) | PD | DLB | p value |  |  |
| --- | --- | --- | --- | --- | --- | --- | --- |
|  |  |  |  |  | LR vs HR | LR vs PD | LR vs DLB |
| Number (M:F) | 37 (26:11) | 82 (65:35) | 84 (44:40) | 16 (8:8) | 0.822<sup>a</sup> | 0.295 | 0.411 |
| Age, years | 63.8 (5.2) | 64.9 (7.6) | 68.8 (9.4) | 78.4 (5.4) | 0.909<sup>b</sup> | 0.008 | <0.001 |
| Education, years | 14.2 (1.9) | 13.5 (2.1) | 13.3 (3.0) | 12.4 (3.9) | 0.530<sup>b</sup> | 0.246 | 0.110 |
| DaT abnormal, % | 3/37 (8.1) | 21/82 (25.6) | 43/43 (100) | 12/12 (100) | 0.028<sup>c</sup> |  |  |
| MIBG abnormal, % | 3/37 (8.1) | 15/82 (18.3) | 41/48 (85.4) | 7/9 (77.8) | 0.178<sup>c</sup> |  |  |
| DaT and/or MIBG abnormal, % | 3/37 (8.1) | 30/82 (36.6) | 62/62 (100) | 15/15 (100) | <0.001<sup>c</sup> |  |  |
| Disease duration, years | NA | NA | 5.9 (4.9) | 3.7 (3.9) |  |  |  |
| MoCA-J | 27.1 (2.4) | 26.7 (2.9) | 24.5 (4.0) | 14.7 (6.9) | 0.782<sup>d</sup> | 0.048 | <0.001 |
| Stroop test part 2 - part 1, sec | 9.7 (4.9) | 12.7 (8.6) | 21.6 (33.7)* | 46.11 (34.70)* | 0.597<sup>d</sup> | 0.140 | 0.004 |
| Line orientation test | 18.2 (2.3) | 17.0 (2.9) | 15.8 (2.9) | 12.9 (3.8)** | 0.100<sup>d</sup> | 0.008 | <0.001 |
| Hoehn and Yahr | 0.0 (0.0) | 0.0 (0.0) | 2.1 (0.9) | 2.8 (1.4) |  |  |  |
| LEDD | 0.0 (0.0) | 0.0 (0.0) | 403.1 (388.3) | 25.0 (77.5) |  |  |  |
| MDS-UPDRS III | 2.2 (2.5) | 4.4 (4.1) | 25.0 (10.3) | 28.9 (22.1) | 0.488<sup>d</sup> | <0.001 | <0.001 |
| Rigidity | 0.2 (0.6) | 0.4 (0.7) | 3.6 (3.1) | 2.6 (2.6) | 0.721<sup>d</sup> | <0.001 | 0.038 |
| Tremor | 0.4 (0.9) | 0.5 (0.9) | 3.8 (4.2) | 2.0 (5.5) | 0.969<sup>d</sup> | <0.001 | 0.886 |
| Bradykinesia | 1.4 (1.7) | 2.3 (2.4) | 12.5 (5.6) | 16.3 (10.8) | 0.296<sup>d</sup> | <0.001 | <0.001 |
| Axial signs | 0.2 (0.6) | 1.2 (1.4) | 5.2 (3.8) | 8.1 (6.2) | 0.126<sup>d</sup> | <0.001 | <0.001 |
| SCOPA-AUT | 1.9 (1.7) | 10.2 (5.1) | 11.6 (8.4) | 13.4 (8.8) | <0.001<sup>d</sup> | <0.001 | <0.001 |
| SAOQ, % | 99.7 (1.1) | 83.2 (25.9) | 67.9 (36.5) | 56.1 (43.6) | 0.009<sup>d</sup> | <0.001 | <0.001 |
| RBDSQ | 0.9 (0.9) | 4.6 (2.8) | 4.0 (2.8) | 4.1 (3.1) | <0.001<sup>d</sup> | <0.001 | <0.001 |
| BDI-II | 2.0 (2.0) | 11.0 (6.9) | 9.8 (6.4) | 10.4 (7.1) | <0.001<sup>d</sup> | <0.001 | <0.001 |
| ESS | 4.8 (2.8) | 9.3 (4.9) | 7.9 (4.8) | 8.8 (6.2) | <0.001<sup>d</sup> | <0.001 | <0.001 |
| PDQ-39 summary index | 1.2 (1.5) | 11.4 (8.9) | 19.4 (15.5) | 24.5 (17.4) | <0.001<sup>d</sup> | <0.001 | <0.001 |
| QUIP | 0.1 (0.4) | 0.8 (1.4) | 0.4 (0.9) | 0.7 (1.7) | 0.004<sup>d</sup> | 0.164 | 0.066 |

PD, Parkinson's disease; DLB, Dementia with Lewy bodies; DaT, dopamine transporter; MIBG, metaiodobenzylguanidine; MoCA-J, the Japanese version of the Montreal Cognitive Assessment; LEDD, Levodopa equivalent daily dose; MDS-UPDRS, Movement Disorder Society-Unified Parkinson's Disease Rating Scale; SCOPA-AUT, the Japanese version of the Scale for Outcomes in Parkinson's disease for Autonomic Symptoms; SAOQ, Self-administered Odor Question; RBDSQ, RBD screening scale; BDI- II, Beck Depression Inventory-Second Edition; ESS, Epworth Sleepiness Scale; PDQ-39, Parkinson's Disease
Questionnaire-39; QUIP, Questionnaire for Impulsive-Compulsive Disorders in Parkinson's disease
\*Two patients with PD and three patients with DLB could not complete the Stroop test
\*\*One patient with DLB could not complete the line orientation test
<sup>a</sup>p values are determined by pairwise comparisons using Fisher's exact test with Benjamini-Hochberg correction
<sup>b</sup>p values determined by a one-way ANOVA with Tukey post-hoc test
<sup>c</sup>p values determined by Fisher's exact test
<sup>d</sup>p values determined by analysis of covariance (ANCOVA) adjusted for age and sex with Tukey's post-hoc test using the Benjamini-Hochberg method
Data represent the mean (standard deviation) or value (%)

The PD-CI group participants were significantly older than the PD-CN group participants. The PD-CI group participants, who were selected based on the MoCA-J scores, had worse line orientation test scores compared with the PD-CN group participants. Compared with the PD-CN group participants, the PD-CI participants had worse RBDSQ, BDI-II, PDQ-39, and QUIP scores, while no significant difference was found in motor function (Supplementary Table 1).

### Plasma biomarkers

Pearson’s correlation analysis between plasma biomarkers and age exhibited a weak correlation for plasma Aβ composite in the PD group, weak correlations for plasma p-tau181 in the low-risk, high-risk, and PD groups, and moderate correlations for plasma NfL in the high-risk and PD groups (Supplementary Figure 1). Therefore, all statistical tests regarding plasma biomarkers were adjusted for age. Considering phenotypic differences of AD between females and males, biomarker values for sex were adjusted as well.[29]

Plasma Aβ composite levels in the DLB group were elevated compared with those in the other groups; however, it was statistically significant only between the DLB and the high-risk groups (Figure 1A). Plasma log_10_ (p-tau181) levels were significantly higher in the PD and DLB groups than in the low– and high-risk groups (Figure 1B). Plasma log_10_ (NfL) levels were significantly higher in the high-risk, PD, and DLB groups than in the low-risk group, with the DLB group exhibiting a pronounced elevation (Figure 1C). Plasma aSyn/Hb ratios were significantly lower in the PD group than in the high-risk and DLB groups. Plasma aSyn/Hb ratios were significantly higher in the DLB group than in the low-risk group (Figure 1D).

**Fig. 1.**
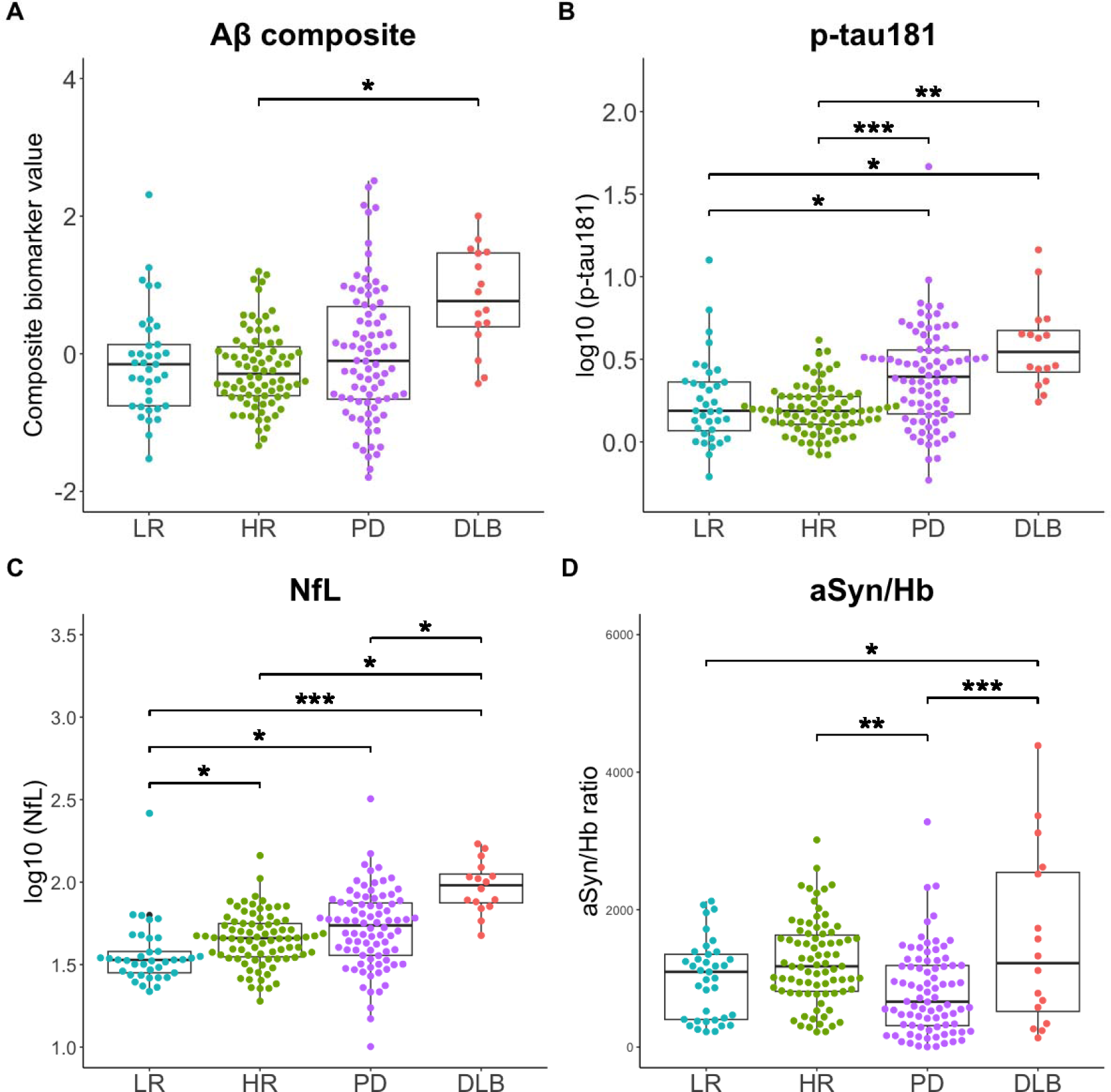
Levels of plasma biomarkers across diagnostic groups. Levels of four plasma biomarkers (Aβ composite (A), p-tau181 (B), NfL (C), and aSyn/Hb (D)) are plotted with individual values and boxplots across diagnostic groups. The plasma p-tau181 and NfL levels were log-transformed with base 10 to approximate a normal distribution. Analysis of covariance (ANCOVA) adjusted for age and sex, followed by Tukey’s tests using the Benjamini–Hochberg method, is used to determine p values visualized with ***p<0.001, **p<0.01, *p<0.05. Aβ composite, a combination biomarker of amyloid-beta precursor protein (APP)_669-711_/amyloid-beta (Aβ)_1-42_ and Aβ_1-40_/Aβ_1-42_ ratios; p-tau181, phosphorylated tau 181; NfL, neurofilament light chain; aSyn/Hb, alpha-synuclein/haemoglobin ratio; LR, low-risk; HR, high-risk; PD, Parkinson’s disease; DLB, dementia with Lewy bodies.

The age-adjusted partial correlation analysis to assess the relationships between plasma biomarkers revealed no correlation between the plasma biomarkers in the low– and high-risk groups (Figure 2A, 2B). In the PD group, Aβ composite and log_10_ (p-tau181) and log_10_ (p-tau181) and log_10_ (NfL) were weakly correlated (Figure 2C). In the DLB group, Aβ composite and log_10_ (p-tau181) were moderately correlated (Figure 2D).

**Fig. 2.**
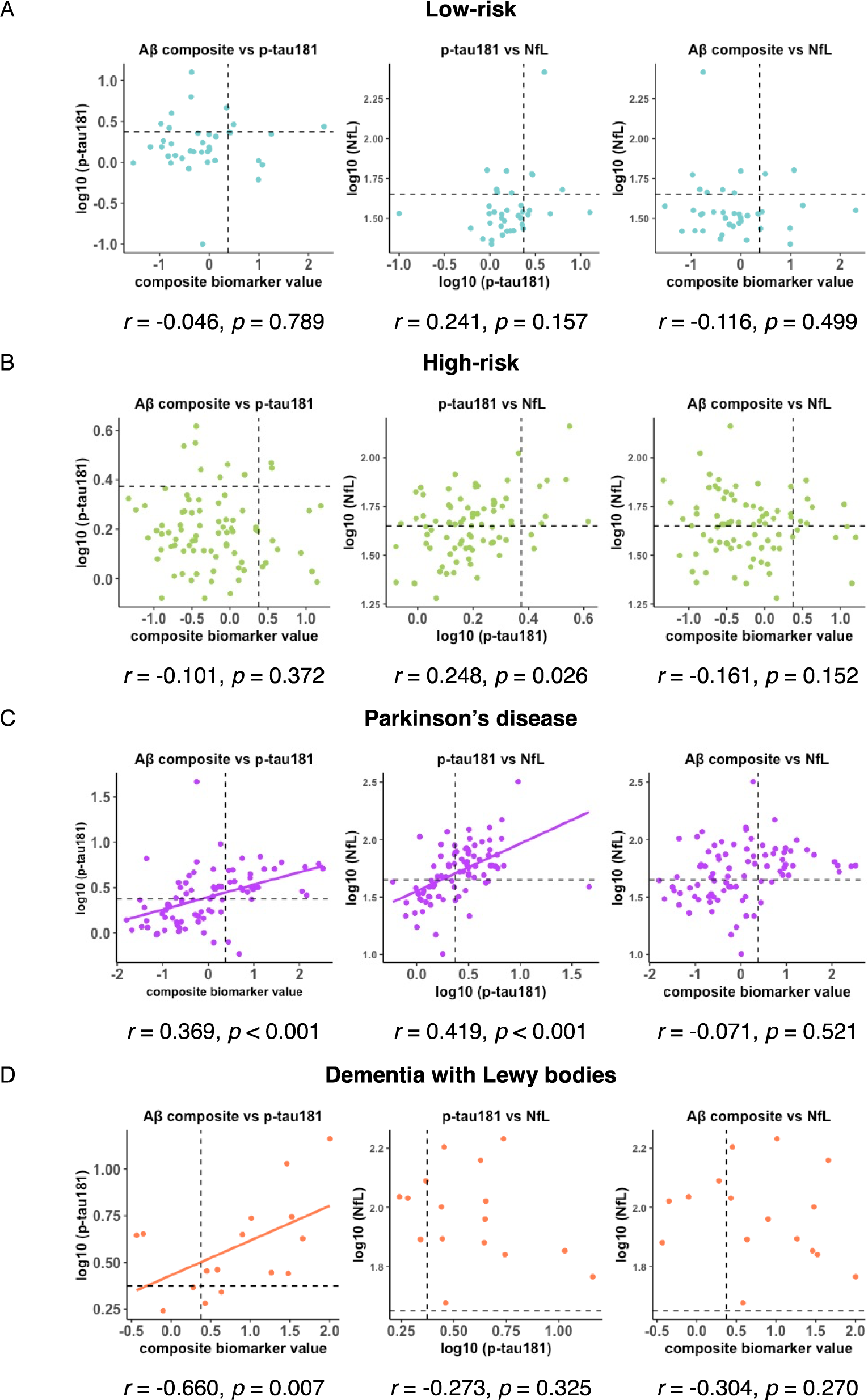
Age-adjusted partial correlation between plasma biomarkers. An age-adjusted Pearson’s partial correlation test among the plasma biomarkers in the low-risk group (A), high-risk group (B), Parkinson’s disease group (C), and dementia with Lewy bodies group (D). Cut-off values for Aβ composite, log_10_ (p-tau181), and log_10_ (NfL) are indicated by dotted lines (Aβ composite, 0.376; log_10_ (p-tau181), 0.374; log_10_ (NfL), 1.65). Aβ composite, combination biomarker of amyloid-beta precursor protein (APP)_669-711_/amyloid-beta (Aβ)_1-42_ and Aβ_1-40_/Aβ_1-42_ ratios; p-tau181, phosphorylated tau 181; NfL, neurofilament light chain; aSyn/Hb, alpha-synuclein/haemoglobin ratio.

The PD-CI group exhibited a significant increase in plasma Aβ composite and log_10_ (p-tau181) levels compared with the PD-CN group (Figure 3A, 3B). Although plasma log_10_ (NfL) levels tended to be higher in the PD-CI group than in the PD-CN group, the difference was not statistically significant (Figure 3C). No significant differences were found in the aSyn/Hb ratio between the PD-CI and PD-CN groups (Figure 3D).

**Fig. 3.**
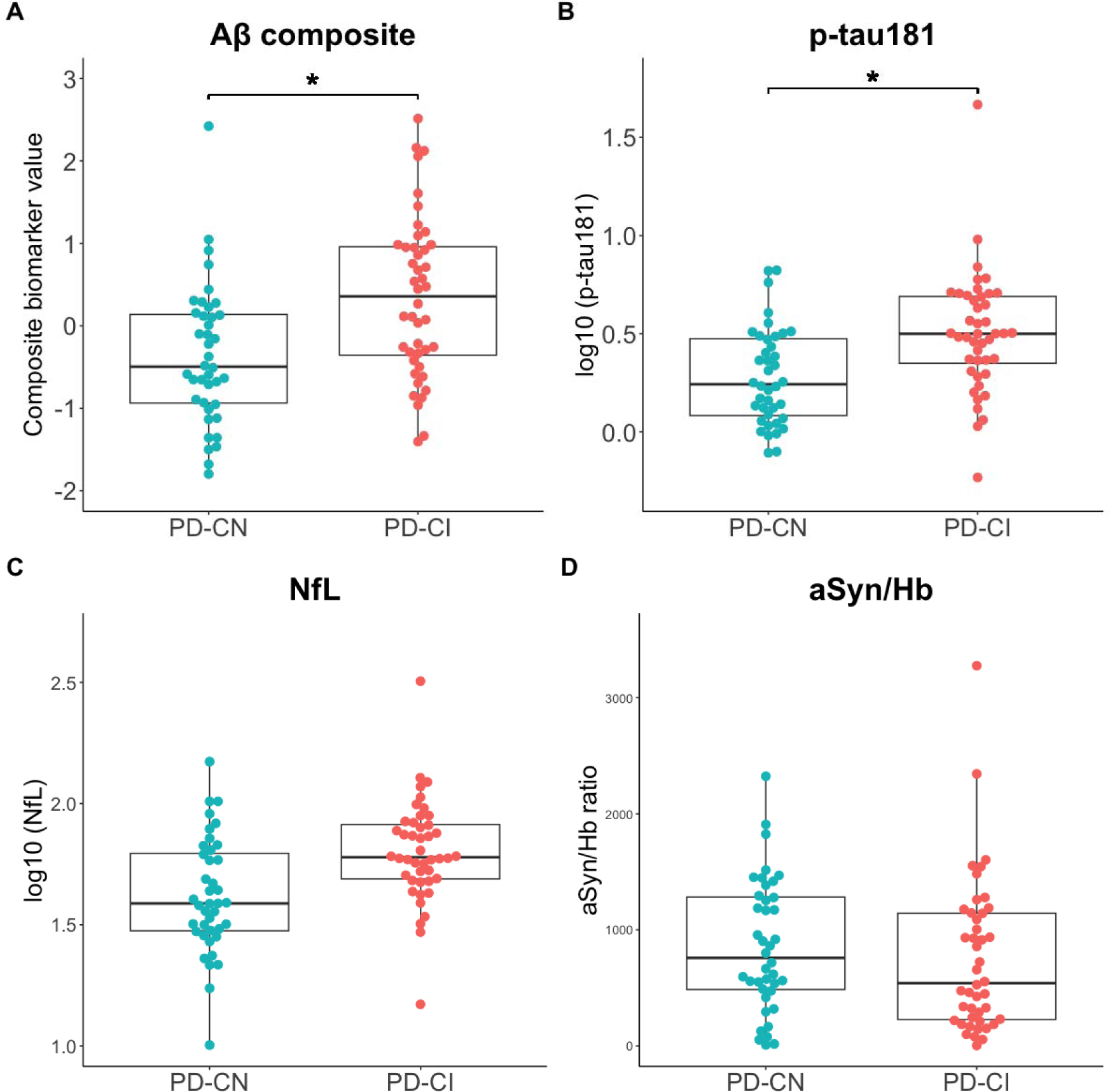
Subgroup analysis of plasma biomarkers by cognitive function in the Parkinson’s disease group. Levels of four plasma biomarkers (Aβ composite (A), p-tau181 (B), NfL (C), and aSyn/Hb (D)) are plotted with individual values and boxplots across diagnostic groups. The patients with PD with MoCA-J ≥26 and <26 are classified as cognitively normal (PD-CN) and cognitively impaired (PD-CI), respectively.[15] The plasma p-tau181 and NfL levels were log-transformed with base 10 to approximate a normal distribution. Analysis of covariance (ANCOVA) adjusted for age and sex is used to determine p values visualized with ***p<0.001, **p<0.01, *p<0.05. Aβ composite, combination biomarker of amyloid-beta precursor protein (APP)_669-711_/amyloid-beta (Aβ)_1-42_ and Aβ_1-40_/Aβ_1-42_ ratios; p-tau181, phosphorylated tau 181; NfL, neurofilament light chain; aSyn/Hb, alpha-synuclein/haemoglobin ratio; PD, Parkinson’s disease; MoCA-J, the Japanese version of the Montreal Cognitive Assessment.

### AT(N) profiles

The proportions of A−T−(N)− and A+T+(N)+ were significantly lower and higher, respectively, in the PD and DLB groups than those in the low-risk group. The proportion of A−T−(N)+ in the high-risk group was significantly higher than that in the low-risk group (Table 2).

**Table 2.**
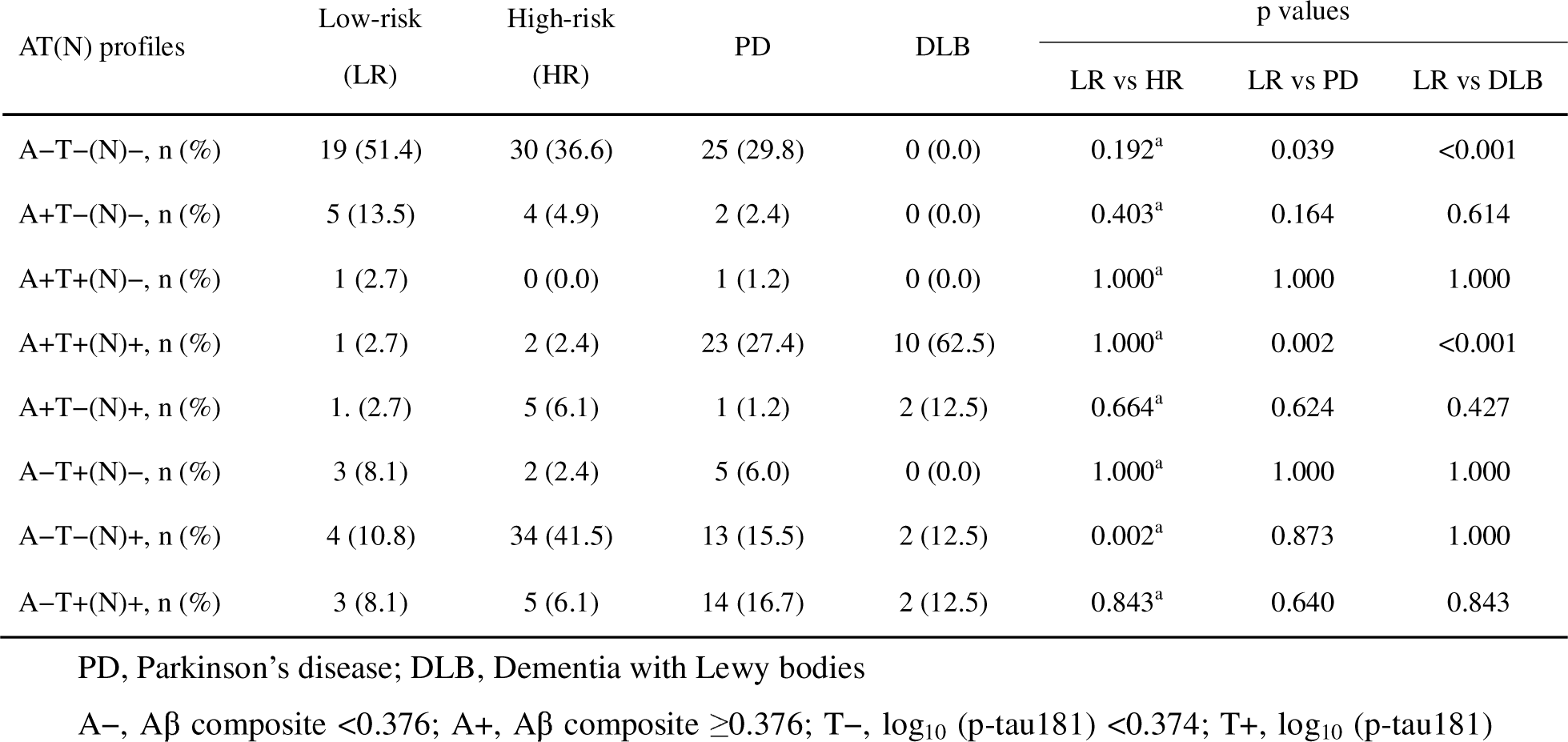
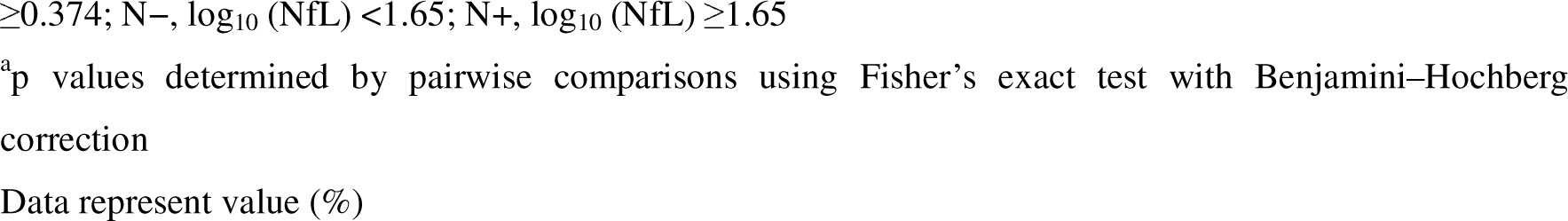
AT(N) profiles of the participants.

| AT(N) profiles | Low-risk<br>(LR) | High-risk<br>(HR) | PD | DLB | p values |  |  |
| --- | --- | --- | --- | --- | --- | --- | --- |
|  |  |  |  |  | LR vs HR | LR vs PD | LR vs DLB |
| A–T–(N)–, n (%) | 19 (51.4) | 30 (36.6) | 25 (29.8) | 0 (0.0) | 0.192<sup>a</sup> | 0.039 | <0.001 |
| A+T–(N)–, n (%) | 5 (13.5) | 4 (4.9) | 2 (2.4) | 0 (0.0) | 0.403<sup>a</sup> | 0.164 | 0.614 |
| A+T+(N)–, n (%) | 1 (2.7) | 0 (0.0) | 1 (1.2) | 0 (0.0) | 1.000<sup>a</sup> | 1.000 | 1.000 |
| A+T+(N)+, n (%) | 1 (2.7) | 2 (2.4) | 23 (27.4) | 10 (62.5) | 1.000<sup>a</sup> | 0.002 | <0.001 |
| A+T–(N)+, n (%) | 1 (2.7) | 5 (6.1) | 1 (1.2) | 2 (12.5) | 0.664<sup>a</sup> | 0.624 | 0.427 |
| A–T+(N)–, n (%) | 3 (8.1) | 2 (2.4) | 5 (6.0) | 0 (0.0) | 1.000<sup>a</sup> | 1.000 | 1.000 |
| A–T–(N)+, n (%) | 4 (10.8) | 34 (41.5) | 13 (15.5) | 2 (12.5) | 0.002<sup>a</sup> | 0.873 | 1.000 |
| A–T+(N)+, n (%) | 3 (8.1) | 5 (6.1) | 14 (16.7) | 2 (12.5) | 0.843<sup>a</sup> | 0.640 | 0.843 |

PD, Parkinson's disease; DLB, Dementia with Lewy bodies
A–, Aβ composite <0.376; A+, Aβ composite ≥ 0.376; T–, log<sub>10</sub> (p-tau181) <0.374; T+, log<sub>10</sub> (p-tau181)
$\geq 0.374$ ; N–, $\log_{10}(\text{NfL}) < 1.65$ ; N+, $\log_{10}(\text{NfL}) \geq 1.65$
<sup>a</sup>p values determined by pairwise comparisons using Fisher's exact test with Benjamini–Hochberg correction
Data represent value (%)

### Differences in clinical features and plasma biomarkers of patients with PD and high-risk individuals across AT(N) profiles

Patients with PD classified as A+ were older and had a shorter educational history, worse MoCA-J scores, and higher plasma log_10_ (p-tau181) levels than those classified as A−. Patients with PD classified as T+ were older and had worse Hoehn and Yahr Scale and MDS-UPDRS III total scores and subscores for bradykinesia and axial signs, and worse SCOPA-AUT, BDI–II, PDQ-39, and QUIP scores than those classified as T−. Patients with PD classified as T+ had higher levels of Aβ composite and log_10_ (NfL) than those classified as T−. Patients with PD classified as N+ were older and had worse scores on the MoCA-J and Hoehn and Yahr Scales, worse MDS-UPDRS III total scores and subscores for bradykinesia and axial signs, worse SCOPA-AUT, BDI-II, PDQ-39, and QUIP scores, and higher levels of plasma Aβ composite and log_10_ (p-tau181) than those classified as N− (Table 3).

**Table 3.** Clinical characteristics of the patients with Parkinson’s disease grouped by A/T/N profiles.

|  | PD |  | p value | PD |  | p value | PD |  | p value |
| --- | --- | --- | --- | --- | --- | --- | --- | --- | --- |
|  | A– | A+ |  | T– | T+ |  | N– | N+ |  |
| Number (M:F) | 57 (31:26) | 27 (13:14) | 0.645<sup>a</sup> | 41 (22:19) | 43 (22:21) | 0.831<sup>a</sup> | 33 (17:16) | 51 (27:24) | 1.000<sup>a</sup> |
| Age, years | 66.0 (9.2) | 74.7 (6.6) | <0.001<sup>b</sup> | 66.0 (9.0) | 71.5 (9.1) | 0.006<sup>b</sup> | 63.2 (9.1) | 72.5 (7.6) | <0.001<sup>b</sup> |
| Education, years | 13.8 (2.6) | 12.2 (3.6) | 0.021<sup>b</sup> | 13.5 (2.8) | 13.0 (3.3) | 0.444<sup>b</sup> | 13.7 (2.4) | 13.0 (3.4) | 0.315<sup>b</sup> |
| DaT abnormal, % | 33/33 (100) | 10/10 (100) |  | 24/24 (100) | 19/19 (100) |  | 18/18 (100) | 25/25 (100) |  |
| MIBG abnormal, % | 28/34 (82.4) | 13/14 (92.9) |  | 20/23 (87.0) | 21/25 (84.0) |  | 17/21 (81.0) | 24/27 (88.9) |  |
| DaT and/or MIBG abnormal, % | 43/43 (100) | 19/19 (100) |  | 32/32 (100) | 30/30 (100) |  | 27/27 (100) | 35/35 (100) |  |
| Disease duration, years | 5.5 (4.1) | 6.7 (6.4) | 0.166<sup>c</sup> | 5.3 (4.3) | 6.5 (5.5) | 0.170<sup>c</sup> | 5.1 (3.6) | 6.4 (5.6) | 0.094<sup>c</sup> |
| MoCA-J | 25.6 (3.1) | 22.2 (4.7) | 0.032<sup>c</sup> | 25.7 (3.3) | 23.4 (4.3) | 0.109<sup>c</sup> | 26.8 (2.4) | 23.0 (4.1) | 0.006<sup>c</sup> |
| Stroop test part 2 - part 1, sec | 17.0 (13.4)* | 32.4 (55.7)* | 0.140<sup>c</sup> | 14.4 (10.5)* | 28.4 (45.2)* | 0.129<sup>c</sup> | 11.7 (8.4) | 28.3 (41.9)* | 0.079<sup>c</sup> |
| Line orientation test | 16.0 (3.2) | 15.3 (2.2) | 0.863<sup>c</sup> | 16.0 (3.4) | 15.6 (2.5) | 0.903<sup>c</sup> | 16.6 (2.6) | 15.3 (3.1) | 0.349<sup>c</sup> |
| Hoehn and Yahr | 2.0 (0.9) | 2.5 (0.9) | 0.075<sup>c</sup> | 1.9 (0.8) | 2.4 (1.0) | 0.018<sup>c</sup> | 1.8 (0.8) | 2.4 (0.9) | 0.013<sup>c</sup> |
| LEDD | 409.6 (427.1) | 389.1 (297.0) | 0.496<sup>c</sup> | 337.9 (306.2) | 465.2 (447.9) | 0.028<sup>c</sup> | 312.5 (281.1) | 461.6 (436.9) | 0.002<sup>c</sup> |
| MDS-UPDRS III | 23.7 (10.7) | 27.7 (8.8) | 0.259<sup>c</sup> | 21.7 (8.9) | 28.2 (10.6) | 0.009<sup>c</sup> | 20.0 (8.6) | 28.3 (10.0) | <0.001<sup>c</sup> |
| Rigidity | 3.6 (2.7) | 3.5 (3.8) | 0.332<sup>c</sup> | 3.3 (2.6) | 3.8 (3.5) | 0.715<sup>c</sup> | 3.1 (2.5) | 3.9 (3.4) | 0.681<sup>c</sup> |
| Tremor | 3.2 (3.4) | 5.0 (5.3) | 0.151<sup>c</sup> | 3.5 (4.0) | 4.0 (4.4) | 0.813<sup>c</sup> | 3.3 (4.0) | 4.0 (4.3) | 0.988<sup>c</sup> |
| Bradykinesia | 12.4 (5.9) | 12.8 (4.8) | 0.501<sup>c</sup> | 11.2 (5.2) | 13.8 (5.7) | 0.014<sup>c</sup> | 10.5 (4.7) | 13.8 (5.7) | <0.001<sup>c</sup> |
| Axial signs | 4.5 (3.9) | 6.5 (3.3) | 0.196<sup>c</sup> | 3.7 (3.0) | 6.5 (4.0) | 0.004<sup>c</sup> | 3.0 (2.7) | 6.6 (3.7) | <0.001<sup>c</sup> |
| SCOPA-AUT | 11.8 (7.7) | 11.0 (9.7) | 0.961<sup>c</sup> | 9.3 (6.6) | 13.7 (9.3) | 0.004<sup>c</sup> | 9.2 (5.4) | 13.1 (9.6) | 0.003<sup>c</sup> |
| SAOQ, % | 68.0 (37.1) | 67.7 (35.9) | 0.902<sup>c</sup> | 68.0 (38.5) | 67.8 (35.0) | 0.911<sup>c</sup> | 66.1 (37.6) | 69.1 (36.1) | 0.426<sup>c</sup> |
| RBDSQ | 4.0 (2.7) | 3.9 (3.0) | 0.634<sup>c</sup> | 3.9 (2.7) | 4.1 (2.9) | 0.908<sup>c</sup> | 3.6 (2.1) | 4.2 (3.1) | 0.598<sup>c</sup> |
| BDI-II | 9.7 (5.8) | 10.0 (7.5) | 0.555<sup>c</sup> | 7.9 (5.1) | 11.5 (7.0) | 0.002<sup>c</sup> | 8.4 (5.5) | 10.7 (6.8) | 0.008<sup>c</sup> |
| ESS | 8.4 (4.5) | 6.9 (5.5) | 0.915<sup>c</sup> | 7.9 (4.5) | 7.86 (5.24) | 0.441<sup>c</sup> | 8.6 (4.3) | 7.5 (5.2) | 0.811<sup>c</sup> |
| PDQ-39 summary index | 19.7 (15.6) | 18.7 (15.6) | 0.900<sup>c</sup> | 15.3 (14.2) | 23.3 (15.9) | 0.005<sup>c</sup> | 14.5 (10.2) | 22.5 (17.6) | 0.001<sup>c</sup> |
| QUIP | 0.4 (0.8) | 0.5 (1.1) | 0.069<sup>c</sup> | 0.2 (0.6) | 0.6 (1.1) | 0.005<sup>c</sup> | 0.2 (0.6) | 0.5 (1.0) | 0.004<sup>c</sup> |
| Aβ composite | -0.53 (0.56) | 1.14 (0.61) | <0.001<sup>c</sup> | -0.57 (0.65) | 0.56 (0.90) | <0.001<sup>c</sup> | -0.55 (0.65) | 0.37 (0.98) | 0.002<sup>c</sup> |
| log<sub>10</sub> (p-Tau181) | 0.33 (0.29) | 0.54 (0.24) | 0.026<sup>c</sup> | 0.16 (0.15) | 0.62 (0.22) | <0.001<sup>c</sup> | 0.22 (0.33) | 0.51 (0.20) | <0.001<sup>c</sup> |
| log<sub>10</sub> (NfL) | 1.67 (0.26) | 1.81 (0.14) | 0.748<sup>c</sup> | 1.60 (0.23) | 1.83 (0.19) | <0.001<sup>c</sup> | 1.48 (0.14) | 1.87 (0.15) | <0.001<sup>c</sup> |
| aSyn, pg/ml | 11665.2 (8920.0) | 9942.2 (8392.1) | 0.625<sup>c</sup> | 12083.8 (10187.0) | 10184.2 (7134.9) | 0.858<sup>c</sup> | 11603.1 (8905.1) | 10793.2 (8706.8) | 0.301<sup>c</sup> |
| aSyn/Hb ratio | 841.7<br>(640.60) | 725.2<br>(570.7) | 0.824<sup>c</sup> | 844.5<br>(704.7) | 765.9<br>(527.9) | 0.935<sup>c</sup> | 793.4<br>(577.3) | 811.3<br>(648.5) | 0.182<sup>c</sup> |

PD, Parkinson's disease; DaT, dopamine transporter; MIBG, metaiodobenzylguanidine; MoCA-J, the Japanese version of the Montreal Cognitive Assessment; LEDD, Levodopa equivalent daily dose; MDS-UPDRS, Movement Disorder Society-Unified Parkinson's Disease Rating Scale; SCOPA-AUT, the Japanese version of the Scale for Outcomes in Parkinson's disease for Autonomic Symptoms; SAOQ, Self-administered Odor Question; RBDSQ, RBD screening scale; BDI-II; Beck Depression Inventory-Second Edition; ESS, Epworth Sleepiness Scale; PDQ-39, Parkinson's Disease Questionnaire-39; QUIP, Questionnaire for Impulsive-Compulsive Disorders in Parkinson's disease; Aβ , amyloid-beta; p-tau181, Phosphorylated tau 181; NfL, Neurofilament light chain; aSyn, alpha-Synuclein A–, Aβ composite <0.376; A+, Aβ composite ≥ 0.376; T–, log<sub>10</sub> (p-tau181) <0.374; T+, log<sub>10</sub> (p-tau181) ≥ 0.374; N–, log<sub>10</sub> (NfL) <1.65; N+, log<sub>10</sub> (NfL) ≥ 1.65
\*One patient with PD could not complete the Stroop test
<sup>a</sup>p values determined by Fisher's exact test
<sup>b</sup>p values determined by Student's *t*-test
<sup>c</sup>p values determined by analysis of covariance (ANCOVA) adjusted for age and sex
Data represent the mean (standard deviation) or value (%)

The age-adjusted partial correlation analysis that assessed the relationships between plasma biomarkers and clinical indices (Supplementary Figure 2) revealed that plasma log_10_ (NfL) was weakly correlated with MDS-UPDRS III bradykinesia and axial signs subscores and the SCOPA-AUT and PDQ-39 scores. In addition, plasma log_10_ (p-tau181) was weakly correlated with the SCOPA-AUT and PDQ-39 scores, while plasma aSyn/Hb ratio was weakly correlated with the MDS-UPDRS III rigidity subscore.

No significant differences in age, cognitive and motor functions, questionnaire survey scores, or plasma biomarkers were found between the high-risk participants classified as A+ and A−. The high-risk participants classified as T+ were significantly older and had worse scores on the Hoehn and Yahr Scale, MDS-UPDRS Part III rigidity subscore, and the RBDSQ, and QUIP scales than those classified as T−. The high-risk participants classified as N+ were significantly older and had a higher rate of MIBG abnormalities than those classified as N− (Table 4). The age-adjusted partial correlation analysis revealed no significant correlations between plasma biomarkers and each clinical score (Supplementary Figure 3).

**Table 4.** Clinical characteristics of the high-risk subjects grouped by A/T/N profiles.

|  | High-risk |  | p value | High-risk |  | p value | High-risk |  | p value |
| --- | --- | --- | --- | --- | --- | --- | --- | --- | --- |
|  | A– | A+ |  | T– | T+ |  | N– | N+ |  |
| Number (M:F) | 71 (44:27) | 11 (9:2) | 0.313<sup>a</sup> | 73 (47:26) | 9 (6:3) | 1.000<sup>a</sup> | 36 (22:14) | 46 (31:15) | 0.644<sup>a</sup> |
| Age, years | 64.3 (7.3) | 68.5 (8.9) | 0.090<sup>b</sup> | 64.1 (7.3) | 71.3 (7.1) | 0.006<sup>b</sup> | 61.3 (7.2) | 67.7 (6.7) | <0.001<sup>b</sup> |
| Education, years | 13.4 (2.1) | 14.5 (2.5) | 0.116<sup>b</sup> | 13.6 (2.0) | 13.1 (2.9) | 0.553<sup>b</sup> | 13.2 (1.7) | 13.7 (2.4) | 0.279<sup>b</sup> |
| DaT SBR average | 6.50 (1.39) | 6.37 (2.12) | 0.854<sup>c</sup> | 6.48 (1.22) | 6.56 (3.06) | 0.547<sup>c</sup> | 6.58 (1.25) | 6.41 (1.67) | 0.876<sup>c</sup> |
| DaT Asymmetry Index | 5.12 (3.60) | 6.31 (7.32) | 0.496<sup>c</sup> | 5.45 (4.36) | 3.88 (2.87) | 0.185<sup>c</sup> | 5.18 (3.46) | 5.37 (4.80) | 0.889<sup>c</sup> |
| DaT abnormal, % | 16/71 (22.5) | 5/11 (45.5) | 0.139<sup>a</sup> | 17/73 (23.3) | 4/9 (44.4) | 0.224<sup>a</sup> | 9/36 (25.0) | 12/46 (26.1) | 1.000<sup>a</sup> |
| MIBG early | 2.95 (0.63) | 2.79 (0.54) | 0.685<sup>c</sup> | 2.92 (0.59) | 3.04 (0.84) | 0.318<sup>c</sup> | 2.99 (0.58) | 2.89 (0.65) | 0.867<sup>c</sup> |
| MIBG delay | 3.16 (0.91) | 2.94 (0.78) | 0.880<sup>c</sup> | 3.14 (0.87) | 3.02 (1.08) | 0.721<sup>c</sup> | 3.32 (0.78) | 2.97 (0.95) | 0.402<sup>c</sup> |
| MIBG washout ratio, % | 22.59 (19.52) | 21.47 (19.55) | 0.291<sup>c</sup> | 21.34 (18.93) | 31.31 (22.13) | 0.658<sup>c</sup> | 16.74 (14.17) | 26.90 (21.81) | 0.322<sup>c</sup> |
| MIBG abnormal, % | 13/71 (18.3) | 2/11 (18.2) | 1.000<sup>a</sup> | 13/73 (17.8) | 2/9 (22.2) | 0.666<sup>a</sup> | 2/36 (5.6) | 13/46 (28.3) | 0.018<sup>a</sup> |
| DaT and/or MIBG abnormal, % | 25/71 (35.2) | 5/11 (45.5) | 0.520<sup>a</sup> | 26/73 (35.6) | 4/9 (44.4) | 0.718<sup>a</sup> | 10/36 (27.8) | 20/46 (43.5) | 0.170<sup>a</sup> |
| MoCA-J | 26.6 (3.1) | 27.4 (1.4) | 0.076<sup>c</sup> | 26.8 (2.9) | 25.6 (2.9) | 0.929<sup>c</sup> | 26.9 (2.2) | 26.4 (3.4) | 0.350<sup>c</sup> |
| Stroop test part 2 - part 1, sec | 12.2 (8.6) | 17.3 (8.0) | 0.554<sup>c</sup> | 12.4 (8.7) | 15.4 (7.6) | 0.928<sup>c</sup> | 10.5 (6.7) | 14.5 (9.5) | 0.546<sup>c</sup> |
| Line orientation test | 17.0 (3.1) | 17.2 (1.7) | 0.494<sup>c</sup> | 17.1 (2.8) | 16.2 (3.4) | 0.929<sup>c</sup> | 17.5 (2.7) | 16.6 (3.0) | 0.917<sup>c</sup> |
| MDS-UPDRS III | 4.4 (4.4) | 4.6 (2.1) | 0.373<sup>c</sup> | 4.0 (3.8) | 7.6 (5.4) | 0.200<sup>c</sup> | 3.3 (3.0) | 5.3 (4.7) | 0.744<sup>c</sup> |
| Rigidity | 0.4 (0.8) | 0.4 (0.5) | 0.298<sup>c</sup> | 0.3 (0.7) | 1.1 (0.9) | 0.012<sup>c</sup> | 0.3 (0.5) | 0.5 (0.8) | 0.726<sup>c</sup> |
| Tremor | 0.4 (1.0) | 0.7 (0.8) | 0.586<sup>c</sup> | 0.5 (1.0) | 0.4 (0.7) | 0.466<sup>c</sup> | 0.3 (0.6) | 0.6 (1.1) | 0.743<sup>c</sup> |
| Bradykinesia | 2.4 (2.5) | 1.9 (1.5) | 0.163<sup>c</sup> | 2.2 (2.3) | 3.6 (3.2) | 0.549<sup>c</sup> | 1.9 (2.1) | 2.7 (2.6) | 0.966<sup>c</sup> |
| Axial signs | 1.2 (1.5) | 1.6 (1.0) | 0.940<sup>c</sup> | 1.1 (1.2) | 2.4 (2.7) | 0.077<sup>c</sup> | 0.9 (1.0) | 1.5 (1.6) | 0.668<sup>c</sup> |
| OSIT-J | 8.8 (2.9) | 8.7 (1.4) | 0.489<sup>c</sup> | 9.0 (2.6) | 7.2 (3.4) | 0.261<sup>c</sup> | 8.9 (3.1) | 8.7 (2.4) | 0.333<sup>c</sup> |
| CVRR rest, % | 3.20 (1.71) | 3.14 (1.46) | 0.871<sup>c</sup> | 3.1 (1.5) | 3.7 (2.7) | 0.135<sup>c</sup> | 3.2 (1.5) | 3.2 (1.8) | 0.596<sup>c</sup> |
| SCOPA-AUT | 9.8 (4.8) | 13.0 (5.7) | 0.060<sup>c</sup> | 10.0 (5.1) | 12.2 (4.2) | 0.174<sup>c</sup> | 9.3 (4.7) | 10.9 (5.3) | 0.117<sup>c</sup> |
| SAOQ, % | 81.8 (27.1) | 92.8 (12.9) | 0.086<sup>c</sup> | 84.8 (24.5) | 70.9 (34.4) | 0.214<sup>c</sup> | 84.0 (25.7) | 82.7 (26.2) | 0.756<sup>c</sup> |
| RBDSQ | 4.6 (2.9) | 4.6 (2.5) | 0.980<sup>c</sup> | 4.4 (2.8) | 6.4 (2.4) | 0.022<sup>c</sup> | 5.2 (2.9) | 4.1 (2.7) | 0.055<sup>c</sup> |
| BDI-II | 10.9 (7.2) | 11.2 (5.1) | 0.646<sup>c</sup> | 11.0 (7.1) | 11.00 (5.7) | 0.771<sup>c</sup> | 12.3 (7.1) | 9.9 (6.6) | 0.224<sup>c</sup> |
| ESS | 9.3 (5.1) | 8.8 (4.1) | 0.986<sup>c</sup> | 9.2 (4.8) | 9.8 (6.1) | 0.233<sup>c</sup> | 10.7 (4.9) | 8.1 (4.6) | 0.124<sup>c</sup> |
| PDQ-39 summary index | 11.7 (9.3) | 9.1 (4.7) | 0.591<sup>c</sup> | 11.3 (9.1) | 12.4 (6.8) | 0.348<sup>c</sup> | 13.5 (11.4) | 9.7 (5.8) | 0.206<sup>c</sup> |
| QUIP | 0.8 (1.4) | 1.0 (1.6) | 0.679<sup>c</sup> | 0.7 (1.1) | 1.9 (2.7) | 0.006<sup>c</sup> | 1.1 (1.7) | 0.6 (1.1) | 0.155<sup>c</sup> |
| Aβ composite | -0.38 (0.41) | 0.78 (0.30) | <0.001<sup>c</sup> | -0.24 (0.57) | -0.08 (0.44) | 0.507<sup>c</sup> | -0.19 (0.56) | -0.25 (0.57) | 0.467<sup>c</sup> |
| log<sub>10</sub> (p-Tau181) | 0.20 (0.14) | 0.18 (0.17) | 0.239<sup>c</sup> | 0.16 (0.11) | 0.48 (0.07) | <0.001<sup>c</sup> | 0.15 (0.12) | 0.22 (0.16) | 0.489<sup>c</sup> |
| log<sub>10</sub> (NFL) | 1.65 (0.16) | 1.67 (0.14) | 0.440<sup>c</sup> | 1.64 (0.15) | 1.77 (0.19) | 0.238<sup>c</sup> | 1.51 (0.09) | 1.76 (0.11) | <0.001<sup>c</sup> |
| aSyn, pg/ml | 17613.0 (9048.4) | 20828.5 (9169.9) | 0.355<sup>c</sup> | 18205.8 (9524.1) | 16734.9 (3907.3) | 0.684<sup>c</sup> | 18774.6 (9038.5) | 17472.9 (9161.5) | 0.489<sup>c</sup> |
| aSyn/Hb ratio | 1213.1<br>(613.1) | 1412.8<br>(561.8) | 0.373<sup>c</sup> | 1244.5<br>(634.1) | 1202.3<br>(333.5) | 0.832<sup>c</sup> | 1279.8<br>(596.9) | 1208.7<br>(619.6) | 0.526<sup>c</sup> |

DaT, dopamine transporter; MIBG, metaiodobenzylguanidine; MoCA-J, the Japanese version of the Montreal Cognitive Assessment; MDS-UPDRS, Movement Disorder Society-Unified Parkinson's Disease Rating Scale; OSIT-J, the odor stick identification test for Japanese; CVRR, coefficient of variation of RR intervals; SCOPA-AUT, the Japanese version of the Scale for Outcomes in Parkinson's disease for Autonomic Symptoms; SAOQ, Self-administered Odor Question; RBDSQ, RBD screening scale; BDI-II, Beck Depression Inventory-Second Edition; ESS, Epworth Sleepiness Scale; PDQ-39, Parkinson's Disease Questionnaire-39; QUIP, Questionnaire for Impulsive-Compulsive Disorders in Parkinson's disease; Aβ , amyloid-beta; p-tau181, Phosphorylated tau 181; NfL, Neurofilament light chain; aSyn, alpha-Synuclein A-, Aβ composite <0.376; A+, Aβ composite ≥ 0.376; T-, log<sub>10</sub> (p-tau181) <0.374; T+, log<sub>10</sub> (p-tau181) ≥ 0.374; N-, log<sub>10</sub> (NfL) <1.65; N+, log<sub>10</sub> (NfL) ≥ 1.65
<sup>a</sup>p values determined by Fisher's exact test
<sup>b</sup>p values determined by Student's *t*-test
<sup>c</sup>p values determined by analysis of covariance (ANCOVA) adjusted for age and sex
Data represent the mean (standard deviation) or value (%)

## DISCUSSION

This study measured and analysed plasma Aβ composite, p-tau181, NfL, and aSyn in patients with PD and DLB and high– and low-risk individuals who were identified in a questionnaire survey on prodromal symptoms of LBD. The results revealed that both PD and DLB groups had increased plasma p-tau181 levels, indicating that comorbid AD neuropathology exists in manifest LBD. In addition, plasma NfL levels were elevated in the high-risk group despite the absence of significant elevation in AD-related plasma biomarker levels such as Aβ composite and p-tau181; thus, plasma NfL levels may reflect aSyn-induced neurodegeneration at even the prodromal phase of LBD.

Previous studies demonstrated that higher plasma Aβ composite levels can predict Aβ burden with an approximately 90% accuracy when using Pittsburgh compound-B (PIB)-amyloid positron emission tomography (PET) as the standard of truth,[6] and higher plasma p-tau181 levels can predict Aβ and tau positivity on PET.[7] In the present study, although plasma Aβ composite levels were higher in the DLB group than in the other groups, they were statistically significant only between the DLB and the high-risk groups, conflicting with the significant increase in plasma p-tau181 levels in both the PD and DLB groups. Although this incongruity may be due to the limited statistical power in the multigroup comparison, previous PET studies reported a substantially low incidence of amyloid deposition in PD without dementia.[30, 31] Another plasma biomarker study reported that the plasma Aβ_1-42_/Aβ_1-40_ ratio was increased in the PD without dementia group compared with healthy controls and decreased in the PD with dementia group compared with the PD without dementia group.[9] Collectively, these findings suggest that amyloid pathology develops concurrently with cognitive decline in LBD and that p-tau biomarkers are more sensitive than Aβ biomarkers in early PD.

However, in our focused analysis on the PD group, both AD-related plasma biomarker (Aβ composite and p-tau181) levels were significantly higher in the PD-CI group than in the PD-CN group. This suggests that comorbid AD neuropathology influences the development of cognitive impairment in PD, consistent with previous cerebrospinal fluid (CSF) studies.[32] In the patients with PD, those classified as A+ had worse MoCA-J scores compared with those classified as A−, and those classified as T+ had worse scores on motor function (MDS-UPDRS III subscores on bradykinesia and axial signs), the SCOPA-AUT, PDQ-39, and QUIP scales compared with those classified as T−. These observations are consistent with a previous CSF study which reported that lower CSF Aβ_1-42_ and higher p-tau were associated with delayed memory recall and motor function, respectively.[33] Our results are also in line with another postmortem study which indicates that nigrostriatal dopaminergic neurodegeneration is possibly initiated by tau pathology independently of aSyn aggregation.[34]

Conversely, the AD-related plasma biomarkers were not elevated in the high-risk group. These results are consistent with those reported in previous studies, namely, that the rate of positive amyloid PET in patients with idiopathic RBD was similar to that in cognitively normal individuals.[35–37] Although these findings indicate that AD-related plasma biomarkers become detectable after the manifestation of motor/cognitive symptoms in LBD, comorbid AD neuropathology may subsist at an undetectable level in the prodromal phase and influence disease progression. Therefore, further longitudinal data analysis must elucidate the role of AD-related plasma biomarkers on motor function and non-motor symptoms in high-risk individuals.

Elevated plasma NfL level is a reliable biomarker of neurodegeneration in various diseases.[8] Although results of previous cross-sectional studies on PD on the correlation between plasma NfL levels and cognitive and motor functions are inconclusive, those of prospective studies consistently demonstrate a correlation between baseline plasma NfL and worsening cognitive and motor functions.[38] Another study reported that higher baseline NfL levels in patients with idiopathic RBD was associated with worsening cognitive, motor, and autonomic functions and a higher risk of phenoconversion.[11] The present study demonstrated that plasma NfL levels were significantly elevated in the PD, DLB, and high-risk groups compared with those in the low-risk group. In patients with PD, those classified as N+ had worse scores on the MoCA-J, Hoehn and Yahr, MDS-UPDRS III, SCOPA-AUT, BDI-II, PDQ-39, and QUIP scales than those classified as N−, suggesting that plasma NfL levels are related to cognitive function, and motor and non-motor symptoms. In the high-risk participants, although no significant differences were observed in cognitive function, or motor or non-motor symptoms between those classified as N+ and N−, those classified as N+ had a higher rate of abnormal MIBG findings than those classified as N-, suggesting that plasma NfL indicates aSyn-induced neurodegeneration, particularly its peripheral involvement, at the prodromal phase.

Previous studies demonstrated that CSF aSyn is decreased in patients with PD.[39] However, results for studies on plasma aSyn levels have been inconsistent, possibly because plasma aSyn levels can be affected by contamination with red blood cells in which aSyn is abundant.[28] In the present study, we attempted to correct aSyn for haemoglobin levels. The plasma aSyn/Hb ratio was significantly decreased in the PD group compared with that in the high-risk and DLB groups and significantly elevated in the DLB group compared with that in the low-risk group. This inconsistent result indicates that plasma aSyn measurement via Simoa may have limitations, and techniques, such as real-time quaking-induced conversion (RT-QUIC), may be necessary.[40]

This study has some limitations. First, the sample size was relatively small, and discrepancies in age and sex ratio among the groups were present. Second, high-risk participants were selected based on a questionnaire survey on prodromal symptoms, and phenoconversion is yet to be confirmed. Third, diagnoses of PD and DLB were based on clinical evaluations rather than neuropathological confirmation. Fourth, AT(N) profile was determined only by plasma biomarkers, and no PET or CSF studies were performed. Therefore, further studies that consider these limitations are needed to verify the findings in our study.

## CONCLUSIONS

Our study demonstrated that comorbid AD neuropathology is present at the symptomatic phase of LBD. In PD, plasma Aβ composite was associated with general cognitive function, plasma p-tau181 with motor function and non-motor symptoms, and plasma NfL with cognitive and motor functions and non-motor symptoms. In addition, the elevated plasma NfL levels in the high-risk group, despite the absence of changes in AD-related plasma biomarkers, suggested the potentiality of plasma NfL as a biomarker to detect aSyn-induced neurodegeneration in the prodromal phase of LBD.

## DECLARATIONS

### Author Contributions

KH: study design and concept, manuscript drafting/revision, data analysis/interpretation, data acquisition, research project execution, and statistical analysis.

MH, YS, DT, TF, TU, TTs, MS, KY, KS, YA, YW, AH, MY, HS, and MW: data acquisition and research project execution.

HT and TTo: biomarker measurements and manuscript revision (intellectual content).

AN and SN: organisation of biomarker measurements and manuscript revision (intellectual content).

MK: study design and concept, research project execution, data analysis/interpretation, and manuscript revision (intellectual content).

All authors critically evaluated the manuscript, and approved the final version of manuscript to be submitted.

## Funding

This study was supported by AMED: Grant Numbers JP20lk0201124, JP20dm0107155, JP22dk0207052, JP22dk0207055, and JP22ae0101077; JSPS KAKENHI: Grant Numbers JP21K19443 and JP23H00420; and the Research Funding for Longevity Sciences (Nos. 19-20 and 22-26) from the National Centre for Geriatrics and Gerontology (NCGG), Japan.

## Acknowledgments

The biomarker assays/analyses in this study were conducted as a part of the BATON (Blood-based Amyloid, Tau, and Other Neuropathological Biomarkers) and STREAM (Stratification of super-Early Alzheimer disease by Multimodal biomarkers) projects, in association with the NCGG Biobank. We thank the researchers and staff who supported the NCGG Biobank and BATON and STREAM projects.

## Competing interests

Masahisa Katsuno receives research grants from Nihon Medi-physics and Sumitomo Pharma.

## Consent for publication

All participants provided their consent for publication.

## Ethics approval

This study conformed to Declaration of Helsinki guidelines and the Ethical Guidelines for Medical and Health Research Involving Human Subjects endorsed by the Japanese government. The study was approved by the Ethics Review Committee of Nagoya University Graduate School of Medicine (Nos. 2016-0238, 2016-0328, 2017-0521, 2021-0240). All participants provided written informed consent for participation in the study.

## Data availability statement

Anonymized data will be made available upon request from a qualified investigator.

## Notes

### Summary of Updates

Corrected citation errors; Unified terminology; Acknowledgments updated.

