## Supplemental Material for "Plasma biomarkers of neurodegeneration in patients with Parkinson’s disease and dementia with Lewy bodies and at-risk individuals of Lewy body disease in the NaT-PROBE cohort"

**Contents:**

Supplementary Table: 1

Supplementary Figures: 3

**Supplementary Table 1. Background characteristics of the patients with Parkinson's disease by cognitive function**

| | PD-CN | PD-CI | p value |
|---|---|---|---|
| Number (M:F) | 40 (22:18) | 44 (22:22) | 0.668[a] |
| Age, years | 64.4 (9.0) | 72.8 (7.8) | <0.001[b] |
| Education, years | 13.6 (2.4) | 12.9 (3.5) | 0.284[b] |
| DaT abnormal, % | 23/23 (100) | 20/20 (100) | 1.000[a] |
| MIBG abnormal, % | 21/26 (80.8) | 20/22 (90.9) | 0.429[a] |
| DaT and/or MIBG abnormal, % | 33/33 (100) | 29/29 (100) | 1.000[a] |
| Disease duration, years | 6.3 (5.0) | 5.5 (4.9) | 0.582[c] |
| MoCA-J | 27.6 (1.3) | 21.7 (3.5) | <0.001[c] |
| Stroop test part 2 - part 1, sec | 12.8 (7.5) | 30.0 (45.2)* | 0.062[c] |
| Line orientation test | 17.2 (2.1) | 14.5 (3.0) | <0.001[c] |
| Hoehn and Yahr | 1.9 (0.8) | 2.3 (1.0) | 0.222[c] |
| LEDD | 423.6 (449.6) | 384.3 (327.1) | 0.654[c] |
| MDS-UPDRS III | 22.6 (9.8) | 27.2 (10.3) | 0.121[c] |
| Rigidity | 3.2 (2.5) | 3.9 (3.5) | 0.701[c] |
| Tremor | 3.6 (4.0) | 3.9 (4.4) | 0.754[c] |
| Bradykinesia | 11.8 (5.4) | 13.2 (5.7) | 0.093[c] |
| Axial signs | 4.0 (3.4) | 6.2 (3.8) | 0.074[c] |
| SCOPA-AUT | 10.5 (7.0) | 12.5 (9.4) | 0.077[c] |
| SAOQ, % | 65.1 (38.3) | 70.4 (35.1) | 0.343[c] |
| RBDSQ | 3.3 (2.1) | 4.6 (3.2) | 0.034[c] |
| BDI-II | 8.8 (5.8) | 10.7 (6.8) | 0.045[c] |
| ESS | 8.0 (4.6) | 7.8 (5.1) | 0.205[c] |
| PDQ-39 summary index | 15.8 (11.5) | 22.7 (17.9) | 0.005[c] |
| QUIP | 0.3 (0.8) | 0.5 (1.0) | 0.038[c] |

Patients with PD with MoCA-J ≥26 and <26 are classified as cognitively normal (PD-CN) and cognitively impaired (PD-CI), respectively

PD, Parkinson's disease; DaT, dopamine transporter; MIBG, metaiodobenzylguanidine; MoCA-J, the Japanese version of the Montreal Cognitive Assessment; LEDD, Levodopa equivalent daily dose; MDS-UPDRS, Movement Disorder Society-Unified Parkinson's Disease Rating Scale; SCOPA-AUT, the Japanese version of the Scale for Outcomes in Parkinson's disease for Autonomic Symptoms; SAOQ, Self-administered Odor Question; RBDSQ, RBD screening scale; BDI-II, Beck Depression Inventory-Second

Edition; ESS, Epworth Sleepiness Scale; PDQ-39, Parkinson's Disease Questionnaire-39; QUIP, Questionnaire for Impulsive-Compulsive Disorders in Parkinson's disease

*Two patients with PD patients could not complete the Stroop test

[a]p values determined by Fisher's exact test

[b]p values determined by Student's *t*-test

[c]p values determined by analysis of covariance (ANCOVA) adjusted for age and sex

Data represent the mean (standard deviation) or value (%)

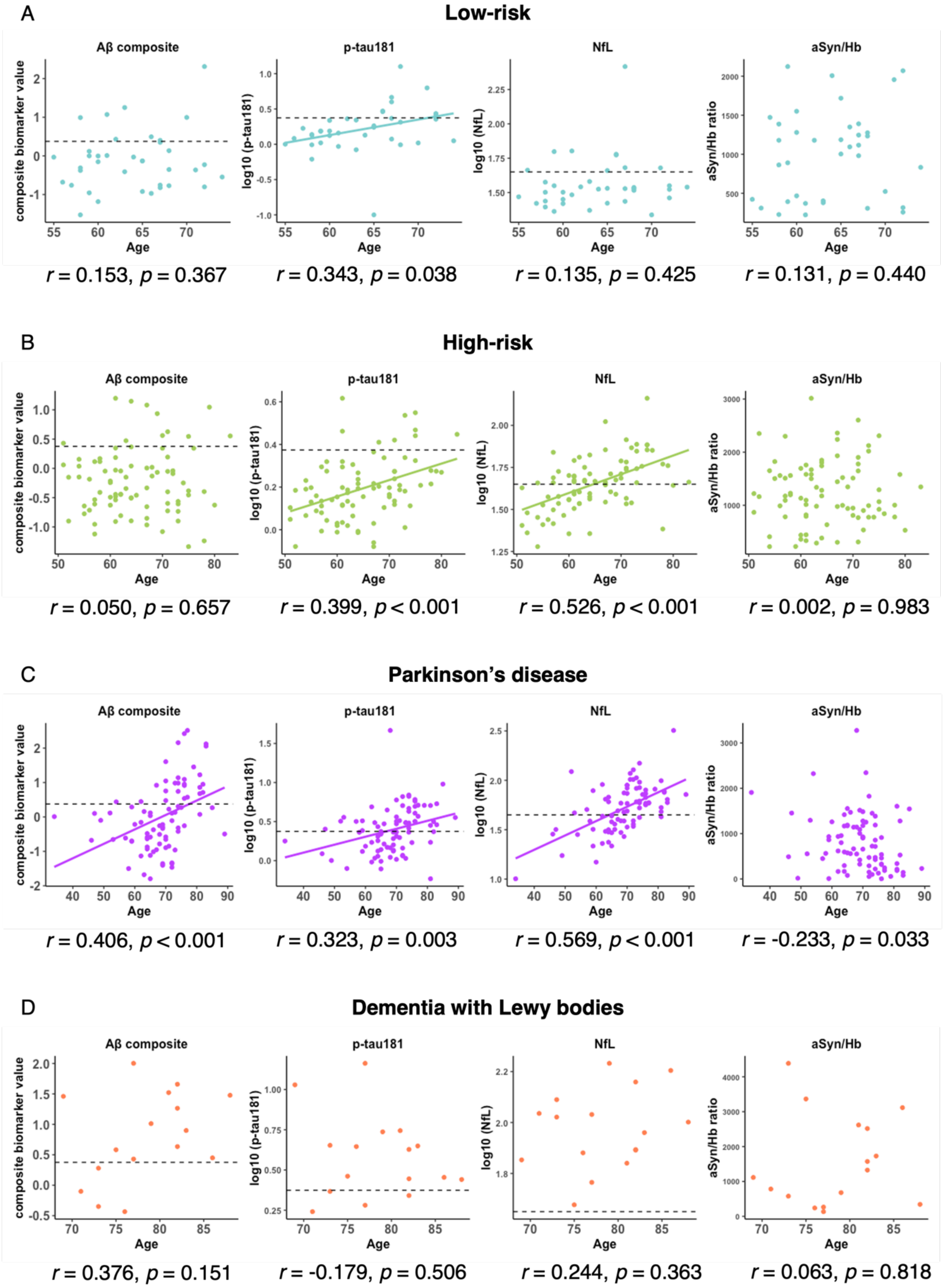


**Supplementary Figure 1. Pearson’s correlation between plasma biomarkers and age**

Pearson’s correlation test between plasma biomarkers and age in the low-risk group (A), high-risk group (B), Parkinson’s disease group (C), and dementia with Lewy bodies group (D). The cut-off values for Aβ composite, $\log_{10}$ (p-tau181), and $\log_{10}$ (NfL) are indicated by dotted lines (Aβ composite, 0.376; $\log_{10}$ (p-tau181), 0.374; $\log_{10}$ (NfL), 1.65).

Aβ composite, combination biomarker of amyloid-beta precursor protein $(APP)_{669-711}$/amyloid-beta $(A\beta)_{1-42}$ and $A\beta_{1-40}/A\beta_{1-42}$ ratios; p-tau181, phosphorylated tau 181; NfL, neurofilament light chain; aSyn/Hb, alpha-synuclein/hemoglobin ratio

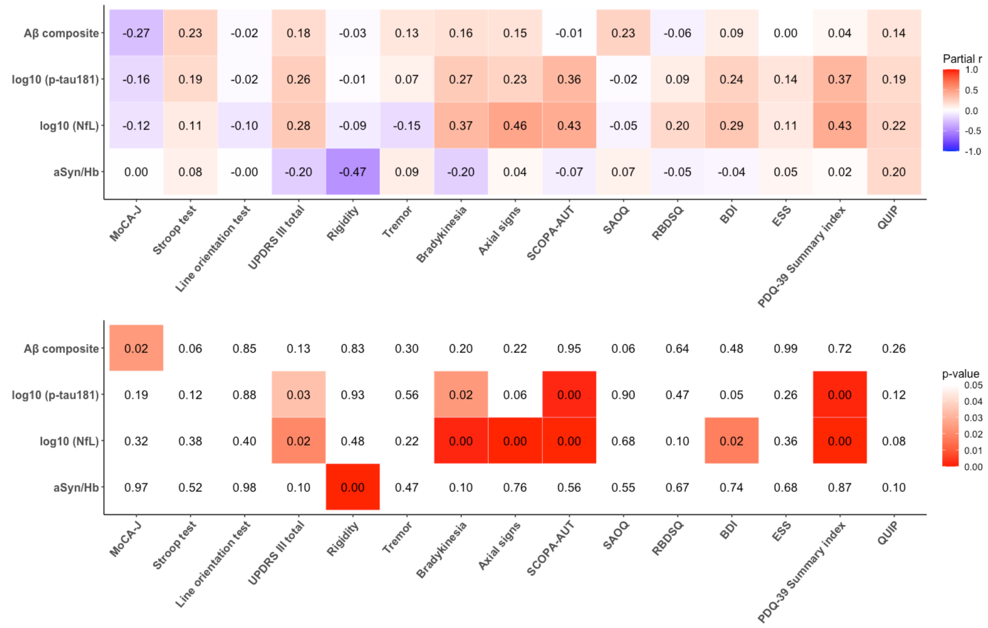


**Supplementary Figure 2. Age-adjusted partial correlation between plasma biomarkers and clinical indices in patients with Parkinson's disease**

Age-adjusted Pearson's partial correlation test between plasma biomarkers and clinical indices in the Parkinson's disease group.

Aβ composite, combination biomarker of amyloid-beta precursor protein (APP)$_{669-711}$/amyloid-beta (Aβ)$_{1-42}$ and Aβ$_{1-40}$/Aβ$_{1-42}$ ratios; p-tau181, phosphorylated tau 181; NfL, neurofilament light chain; aSyn/Hb, alpha-synuclein/hemoglobin ratio; MoCA-J, the Japanese version of the Montreal Cognitive Assessment; UPDRS, Movement Disorder Society-Unified Parkinson's Disease Rating Scale; SCOPA-AUT, the Japanese version of the Scale for Outcomes in Parkinson's disease for Autonomic Symptoms; SAOQ, Self-administered Odor Question; RBDSQ, RBD screening scale; BDI-II, Beck Depression Inventory-Second Edition; ESS, Epworth Sleepiness Scale; PDQ-39, Parkinson's Disease Questionnaire-39; QUIP, Questionnaire for Impulsive-Compulsive Disorders in Parkinson's disease

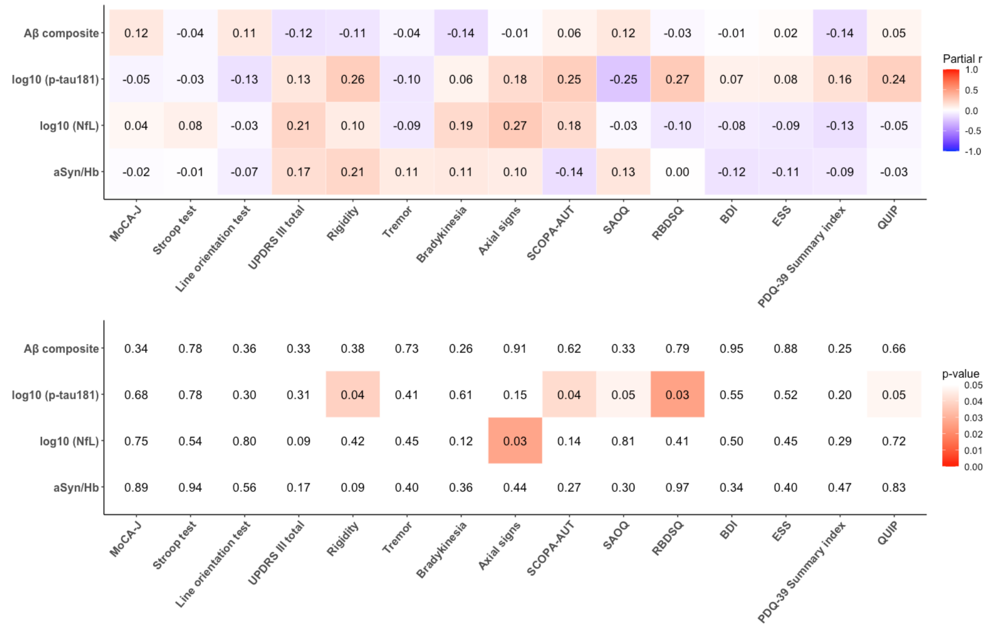


**Supplementary Figure 3. Age-adjusted partial correlation between plasma biomarkers and clinical indices in high-risk individuals**

Age-adjusted Pearson's partial correlation test between plasma biomarkers and clinical indices in the high-risk group.

Aβ composite, combination biomarker of amyloid-beta precursor protein $(APP)_{669-711}$/amyloid-beta $(A\beta)_{1-42}$ and $A\beta_{1-40}/A\beta_{1-42}$ ratios; p-tau181, phosphorylated tau 181; NfL, neurofilament light chain; aSyn/Hb, alpha-synuclein/hemoglobin ratio; MoCA-J, the Japanese version of the Montreal Cognitive Assessment; UPDRS, Movement Disorder Society-Unified Parkinson's Disease Rating Scale; SCOPA-AUT, the Japanese version of the Scale for Outcomes in Parkinson's disease for Autonomic Symptoms; SAOQ, Self-administered Odor Question; RBDSQ, RBD screening scale; BDI-II, Beck Depression Inventory-Second Edition; ESS, Epworth Sleepiness Scale; PDQ-39, Parkinson's Disease Questionnaire-39; QUIP, Questionnaire for Impulsive-Compulsive Disorders in Parkinson's disease
